# High-throughput SARS-CoV-2 and host genome sequencing from single nasopharyngeal swabs

**DOI:** 10.1101/2020.07.27.20163147

**Authors:** J. E. Gorzynski, H. N. De Jong, D. Amar, C. Hughes, A. Ioannidis, R. Bierman, D. Liu, Y. Tanigawa, A. L. Kistler, J. Kamm, J. Kim, L. Cappello, N. F. Neff, S. Rubinacci, O. Delaneau, M. J. Shoura, K. Seo, A. Kirillova, A. Raja, S. Sutton, C. Huang, M. K. Sahoo, K. C. Mallempati, G. Montero-Martin, K. Osoegawa, N. Watson, N. Hammond, R. Joshi, M. A. Fernández-Viña, J. W. Christle, M.T. Wheeler, P. Febbo, K. Farh, G. P. Schroth, F. DeSouza, J. Palacios, J. Salzman, B. A. Pinsky, M. A. Rivas, C.D. Bustamante, E. A. Ashley, V. N. Parikh

## Abstract

During COVID19 and other viral pandemics, rapid generation of host and pathogen genomic data is critical to tracking infection and informing therapies. There is an urgent need for efficient approaches to this data generation at scale. We have developed a scalable, high throughput approach to generate high fidelity low pass whole genome and HLA sequencing, viral genomes, and representation of human transcriptome from single nasopharyngeal swabs of COVID19 patients.

## Main Text

Respiratory virus pandemics, most recently COVID-19 caused by SARS-CoV-2, have caused devastating loss of life and crippled health care systems worldwide. Our public health response to and recovery from these catastrophes depends on the speed and agility with which we generate both viral and host sequencing data. In recent pandemics, viral sequencing has been crucial, but limited by low throughput and high expense. Collection of concomitant host genomics can inform familial relationship tracking, examination of underlying genetic risk, and identify ancestries at elevated risk, but has been encumbered by need for multiple sampling.^1^ Therefore, there is an urgent need for protocols to produce these data in real time and at scale. Here, we describe a method to achieve simultaneous viral and host sequencing from single SARS-CoV-2 diagnostic nasopharyngeal swab residuals **(Figure 1)**. We use low-pass whole host genome sequencing as an alternative to array-based genotyping to provide rich information for trait mapping at scale^2,3^ and demonstrate that our method regularly provides DNA of sufficient quality for host genome, and HLA sequencing. Further, we present a high-throughput RNAseq workflow for sequencing full viral genomes and human transcriptome reads from hundreds of samples concomitantly. Finally, we describe how this method creates a strong multi-omic foundation for data integration and sharing across global institutions.

**Figure 1.**
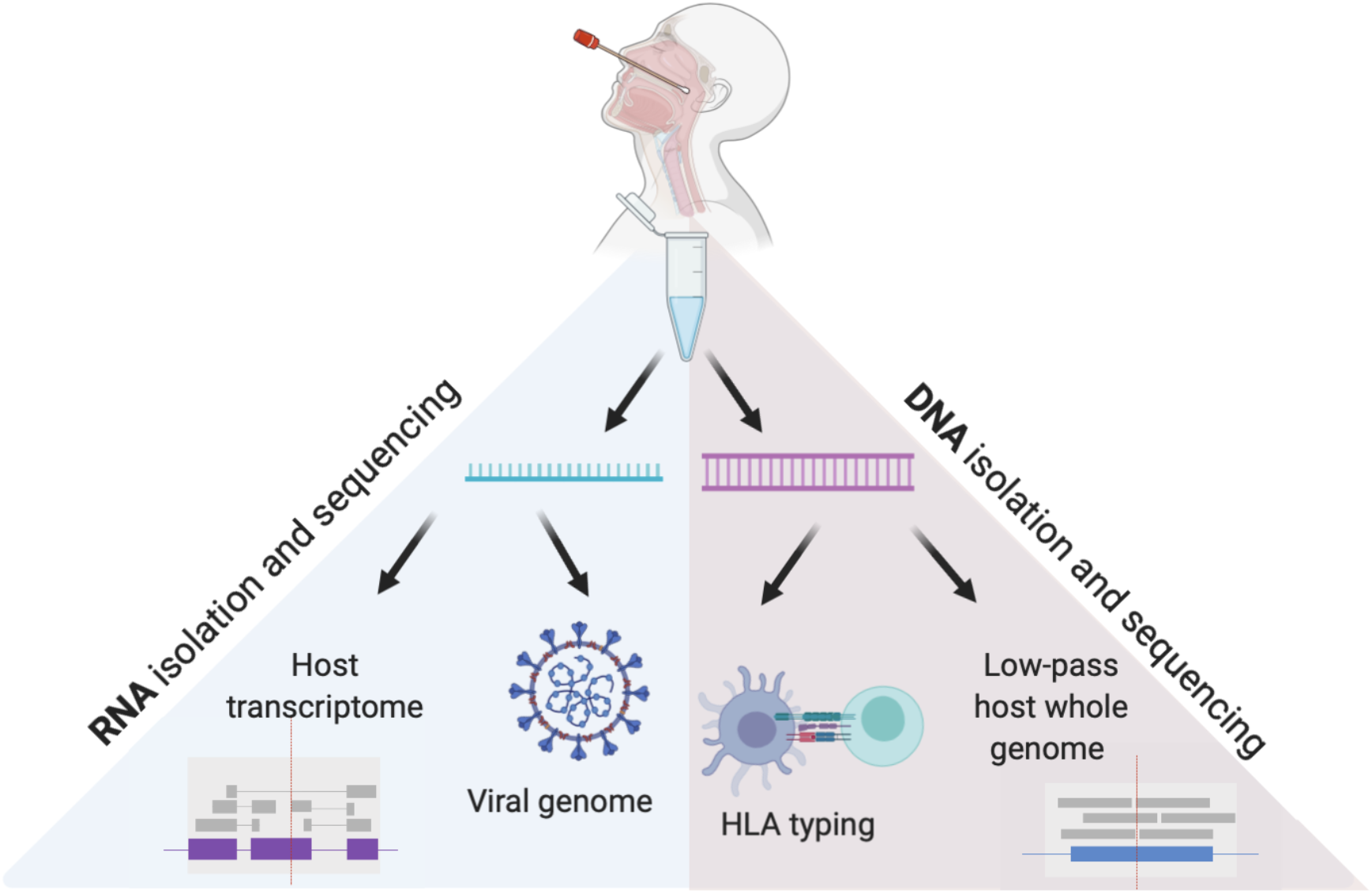
Viral and Host genomes and transcriptomes from a single nasopharyngeal swab. This method allows for independent RNA and DNA isolation from nasopharyngeal swab VTM, enabling viral genome sequencing, detection of host transcriptome, low pass host genome sequencing and HLA sequencing in high throughput.

In this study, residual viral transport media (VTM) from SARS-CoV-2 clinical diagnostic tests were collected. Institutional Review Board approval for anonymous sequencing of host and viral genomics was obtained from the Stanford University School of Medicine IRB. Positive samples are those that had a crossing threshold (CT) of 40 cycles or less on the RT-qPCR diagnostic tests used at Stanford Health Care clinical laboratory.^4^ Virus was inactivated by diluting each sample with one-third volume lysis buffer. Using silica membrane/ethanol nucleotide extraction protocols, we recovered RNA (mean mass: 112ng, 0-1100ng) and DNA (mean mass: 200ng, 0-4700ng) from aliquots of VTM. Seventy percent of samples yielded a total DNA mass greater than 100ng, providing enough input material for many high depth whole genome sequencing protocols.

Multiplexed shotgun sequencing libraries were prepared for RNA and DNA. Purified genomic DNA or cDNA was enzymatically fragmented and unique index adapters were added to the ends by PCR. Fluorometric quantification and capillary electrophoresis fragment size analysis determined the molarity of samples. Samples were pooled at equimolar ratios in batches of 160. For DNA libraries, prior to high-throughput sequencing, low pass sequencing determined sample representation within the pool. Samples were re-pooled if necessary to achieve a balanced library. Pools of 160 genomic DNA or cDNA samples underwent paired-end sequencing by synthesis (2×150bp) using an Illumina NovaSeq and S4 (DNA) or S2 (RNA) flow cell (300 cycles). 117 samples from which more than 2ng/ul of DNA were recovered underwent human leukocyte antigen (HLA) sequencing using at least 20 ng of total sample. In brief, libraries were sequenced for 11 classical HLA genes prepared from batched host genomic DNA samples. Sequences were assembled and assigned to HLA genotypes and used to generate reports for HLA genotypes, HLA serotypes, and imputed HLA haplotypes.

We aligned and called host genomic variants using methods for low pass sequencing ^2,5,6^ followed by imputation with GLIMPSE.^3^ Alignment revealed a mean of 98±3.6% DNA reads mapped to the host genome (GRCh38 assembly), with an average of 61% of the genome covered at 1X, 52% covered at 2X, and 30% covered at 3X. The autosomal mean of medians was 2.21±0.58X, median 2.16X (**Figure 2A**). Variant calling and imputation yielded high fidelity data, as demonstrated by ancestry classification consistent with reference genomes (**Figure 2B**). This also enabled confirmation of six blindly duplicated samples and 2 pairings of first-degree relatives through kinship analysis (**Figure 2C**). Together, these analyses demonstrate the robustness and internal reproducibility of host genome sequencing, variant calling and genotype imputation. We recovered interpretable HLA sequencing from 85% of 116 samples attempted. The remaining samples showed either no amplification of HLA sequences or non-specific amplification. To understand the reason for sample failure, we tested the hypothesis that failed samples had DNA concentration too low to amplify. We found that there was no correlation between DNA concentration and detection (p=0.07).

**Figure 2.**
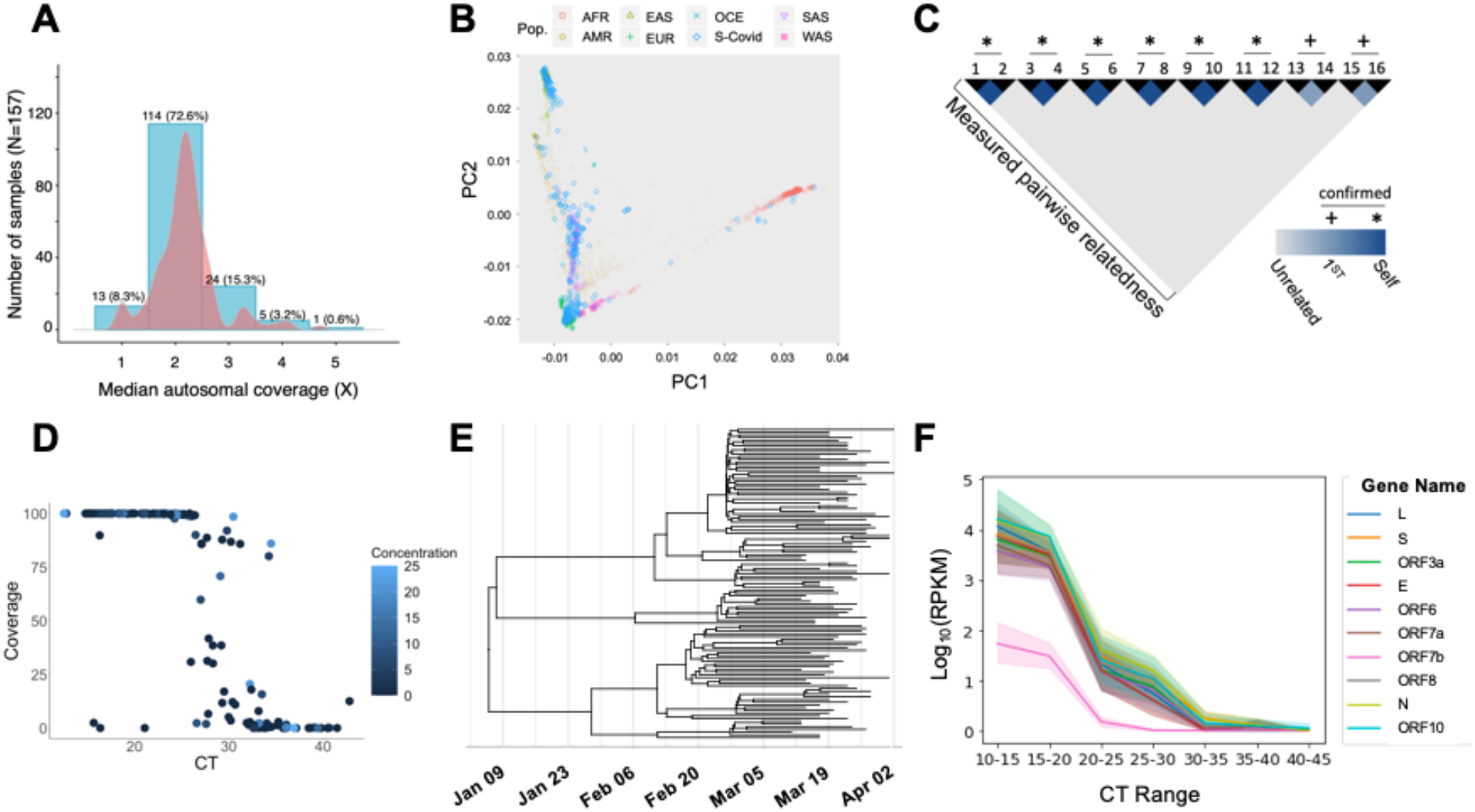
High fidelity, rapid throughput SARS-CoV-2 genome, transcriptome and low pass whole genome sequencing from single nasopharyngeal swabs. **(A)** For low pass genomes, an average of 61% of the human genome was covered at 1X, 52% covered at 2X, and 30% covered at 3X.The autosomal mean of median coverages was 2.21±0.58. **(B)** Principal components analysis shows low pass variant calling and imputation reliably predict ancestry in COVID19 samples (blue diamonds) as compared to reference genomes (ancestries as indicated in legend: AFR - African, AMR - Native American, EAS - East Asian, EUR - European, OCE - Oceanian, SAS - South Asian, WAS - West Asian). **(C)** Blinded duplicates and first degree relatives are predicted by variant calling and imputation of low pass host genomes. **(D)** Percent viral genome covered at 10X at different CT values. The large majority of samples with diagnostic CT number less than 25 yielded >95% of the viral genome using this method. **(E)** Phylogenetic tree of 114 whole viral genomic consensus sequences sampled in March-April 2020. The estimated phylogenetic tree is the maximum clade credibility tree obtained with BEAST 2^15^ using a fixed mutation rate of 1.04×10^-3^ per base per year, the Coalescent Extended Bayesian Skyline prior^16^, and the HKY substitution model^17^. **(F)** RPKMs for individual SARS-CoV-2 genes were averaged over samples with similar CT values. All gene expression increases with viral load (CT range), and all have concordant normalized abundances, with the exception of ORF7b.

RNA sequence was aligned to human transcriptome Hg19, SARS-CoV-2 (MN908947.3 (GenBank)/NC_045512.2 (RefSeq)) and 40 other human viral pathogens (**Supplemental Table 1**) using STAR, kraken2 plus minimap2, and Illumina BaseSpace DRAGEN Pathogen Detection, respectively. RNA sequencing revealed 12.4±22.7% aligned to the SARS-CoV-2 genome. The DRAGEN pipeline identified none of the 160 samples to have 5X coverage at more than 7% of any other viral pathogen genome, indicating no significant co-infections were observed among these samples. On average, 3.9±10.3% of reads aligned to the human genome.

Fifty-two percent of samples had at least 10X coverage at >95% of the SARS-CoV-2 genome. We show that CT number from the Stanford clinical RT-PCR test correlates with viral genome coverage, as well, with a steep drop off in viral genome coverage at CT values of 26 and higher (R^2^=0.61, p=0.0001, **Figure 2D**). Greater than 95% of the genome was covered in 95.3% samples with a diagnostic CT value equal or less than 25. In samples with a CT less than 30, 77% yielded >95% 10X of the viral genome. Three outliers with low CT numbers (high viral load) but low viral genome coverage had low RNA yield from extraction, though overall, total RNA yield did not correlate with viral coverage (R^2^=0.004, p=0.39, **Figure 2D**). Aligned human reads were not inversely correlated with viral genome coverage (R^2^=0.0001, p=0.9), indicating that drop out of viral genome coverage is likely due to other factors, sample contamination or degradation. However, as only three samples with CT numbers below 25 did not yield full viral sequences, we recommend proceeding to library preparation regardless of RNA yield. Using the consensus sequences derived from the initial cohort reported here as well as samples collected later in March 2020, we created a phylogenetic tree, which allows critical public health phylodynamic tracking (**Figure 2E**). Further, we demonstrate high correlation between CT range and detection of all known SARS-CoV-2 genes (**Figure 2F**, R^2^= 0.50, p=5.2e-262). To distinguish reads from the viral genome versus the viral transcriptome (called “sub-genomic” or sgRNA), we also quantified reads containing splicing junctions (which exist solely in sgRNA) and found the same relationship (**Supplemental Figure 1**).

Here we demonstrate that a single nasopharyngeal swab can reveal substantial host *and* viral genomic information in a high-throughput manner that will facilitate public health pandemic tracking and research into the mechanisms underlying virus-host interactions. Certainly, nasopharyngeal swabs have previously been used to perform whole viral genome sequencing of respiratory viruses in low throughput.^7,8,9^ Our method accelerates this discovery both in terms of time and number of subjects sequenced: compared to these reports, we show a comparable rate of viral genomic coverage with the capability of studying at least 10 times the number of samples in a single sequencing run. Although our initial swab collection did not reveal any viral co-infections, especially as the current pandemic enters the regular flu and cold season, our method allows for acceleration of metagenomics analysis.^9,10^ Further, advances in low pass genome calling allow the same nasopharyngeal swab to be used to gather a wealth of human genomics data and in many cases yielded enough DNA for deep sequencing of HLA type, which is a critical component of the host immunomic response. ^2,3^

Future infectious disease outbreaks will inevitably occur, and the strategy we describe is applicable for collection of host and viral genomic information from any respiratory virus in laboratories around the world. Perhaps the most critical application of this workflow is that it enables the rapid development of large scale, multicenter/global host and viral multi-omic data repositories. Global data repositories have been critical to advancing research in the current pandemic and prior. For example, just six months into the SARS-CoV-2 pandemic, nearly 70,000 submissions to the Global Initiative for Sharing All Influenza Data (GISAID) allowed unprecedented tracking of viral mutagenesis and outbreaks.^11^ With the method we propose here, the same number of viral genomes could be produced by less than 100 sequencing centers within weeks, along with matched host genome, transcriptome and HLA typing. These findings could be easily incorporated with data abstracted from the electronic health care record (as is being accomplished with increasing speed^12,13^), and mobile digital reporting platforms^14^ (**Supplemental Figure 2**). The methods described here represent a crucial scaffold for the integration of these complex inputs to centralized data repositories, enabling unprecedented rapidity of the discovery and implementation necessary to overcome these devastating pandemics.

## Methods

### Sample Collection and diagnostics

Residual VTM from SARS-CoV-2 positive nasopharyngeal swabs collected during clinical assessment of asymptomatic and symptomatic patients at Stanford Healthcare were used. RT-qPCR targeting the *envelope* gene or ORF1ab were used to detect infection.. 152 samples with detectable SARS-CoV-2 RNA and 8 with undetectable virus by RT-qPCR were included.

### Nucleic Acid Extraction

DNA; host genomic DNA was extracted from 200ul of VTM inoculated with nasopharyngeal swabs. Using a modified Qiagen DNEASY blood and tissue kit protocol and quantified using fluorometric readings(Supplemental Material, DNA Extraction). Total RNA was extracted from 200ul of VTM using a modified Ambion MirVana mRNA kit protocol (Supplemental Material, RNA extraction) and quantified using fluorometric readings.

### Host gDNA library preparation and sequencing

Using 1-10ng of host gDNA, the Illumina Nextera Flex library preparation was performed according to manufacturer’s protocol (Supplemental Material, DNA Library prep). To allow for multiplexing, gDNA was barcoded using IDT-ILMN Nextera DNA UD Indices, a set of 10bp index adapters from Illumina. Indexed samples were diluted to 4nM, pooled, and analyzed on an Agilent TapeStation to ensure the mean DNA fragment size was ~300bp. Pooling and library quality was further assessed by sequencing the pool using a V3 MiSeq flow cell. 160 samples were pooled and sequenced for 76 cycles, paired end reads. For the purpose of QC, ~ 50 million reads were obtained and Q30 was determined to be >92%. If needed the pool was normalized (balanced) to ensure equal representation of each sample. The library was then sequenced on an Illumina NovaSeq 6000 using an S4 300 cycle flow cell.

### Viral RNA library preparation and sequencing

After extraction, RNA acquired from 100 ul nasal swab media were incubated with recombinant RNAse-free DNase (Qiagen, Inc.) per manufacturer’s instructions for 15 minutes, followed by SPRI bead (GE Healthcare) purification to remove residual DNA remaining in each sample. A fixed volume (5uL) of the resulting RNA from each sample, together with a fixed mass (25pg) of the External RNA Controls Consortium RNA spike-in mix (ERCC RNA spike-in mix, Thermo Fisher), served as input for SARS-CoV-2 metatranscriptomic next generation sequencing (mNGS) library preparation (dx.doi.org/10.17504/protocols.io.beshjeb6; a modification of Deng et al. (2020)^18^). An incubation step with 1:10 dilution of FastSelect (Qiagen) reagent was included between the RNA fragmentation and first strand synthesis steps of the library prep to deplete highly abundant host rRNA sequences present in each sample. Equimolar pools (n=160-384 samples) of the resulting individual dual-barcoded library preps were subjected to paired-end 2 × 150bp sequence analysis on an Illumina NovaSeq 6000 (S2 or equivalent flow cell) to yield approximately 50 million reads per sample.

### Viral Genome Alignment

Metatranscriptomic FASTQ sequences were aligned to the SARS-CoV-2 reference genome NC_045512.2 using minimap2^19^. Non-SARS-CoV-2 reads were filtered out with Kraken2^20^, using an index of human and viral genomes in RefSeq (index downloaded from https://genexa.ch/sars2-bioinformatics-resources/). Spiked primers for viral enrichment were trimmed from the ends of short reads using ivar^21^. Finally, a pileup of the aligned reads was generated with samtools^22^, and consensus genomes were called with ivar. The full pipeline used is publicly available on Github (https://github.com/czbiohub/sc2-illumina-pipeline).

For multiple viral genome alignment, FASTQ files were input into the Illumina BaseSpace DRAGEN Pathogen Detection software with the following parameters: Somatic small variant base caller, k-mers generated from reference (SARSCoV2_NC_045512.2), Minimum depth 10, minimum allele frequency 0.5, virus detected threshold 5% of genome at 5X coverage.

### Quantification of total Viral RNA and sgRNA

RNA mapping was performed using STAR run against a combined index of grch38, SARS-CoV2, and ERCC spike ins. STAR parameters were chosen to avoid bias towards GTAG eukaryotic splice signatures for both the viral RNA and sgRNA analyses ^23^. The STAR parameters used are included in **Supplemental Table 2**. Reads per COVID gene were collected from the ReadsPerGene STAR output file, and the total mappable reads were collected from the Log.final files. Gene lengths were calculated using the annotated start and stop positions in the annotation file (https://ftp.ncbi.nlm.nih.gov/genomes/all/GCF/009/858/895/GCF_009858895.2_ASM985889v3_GCF_009858895.2_ASM985889v3_genomic.gff.gz). Read counts, gene length, and total mappable reads were together used to calculate the RPKM per gene within each sample.

The spliced RNA junction software SICILIAN^24^ was used to sensitively and accurately identify leader-body junctions within each sample. Modifications to standard SICILIAN protocol were to the combined index and STAR mapping parameters described above (**Supplemental Table 2**).^23^

### Host HLA Sequencing

Host genomic DNA samples ranging from 22-75ng were batched in sets of 46 plus one positive and one negative control. AllType™ FASTplex™ NGS Assay kits (One Lambda, A Thermo Fisher Scientific Brand, Canoga Park, CA) were used to prepare DNA sequencing libraries for 11 classical HLA genes (*HLA-A, HLA-C, HLA-B, HLA-DRB3, HLA-DRB4, HLA-DRB5, HLA-DRB1, HLA-DQA1, HLA-DQB1, HLA-DPA1*, and *HLA-DPB1*). As the success of the DNA sequencing is dependent on the initial target amplification and the subsequent library preparation, the following changes were made to the manufacturer’s protocol: 1) increased input DNA volume to 8.6 μl while maintaining the manufacturer’s recommended multiplex PCR protocol per sample; 2) eluted DNA in 12 μl of suspension buffer after the initial amplicon purification, and proceeded to the library preparation without normalization process; 3) increased the number of thermal cycles to 17 in the final DNA library amplification; 4) eluted DNA fragments in 22 μl for DNA sequencing. 500 μl of 1.3 pM DNA sequencing library was loaded into a MiniSeq Mid Output Kit (300-cycles) (FC-420-1004), and sequenced using MiniSeq DNA sequencer (Illumina Inc., San Diego, CA). Fastq files were automatically imported into the TypeStream Visual NGS Analysis Software Version 2.0 upon the completion of DNA sequencing, and bioinformatically processed for DNA sequence assembly and HLA genotype assignments with IPD IMGT/HLA Database release version 3.39.0.^25^ We modified the software setting so that a maximum of 1.5 million sequences or 750,000 paired-end sequences are used for the sequence assembly and HLA allele assignments. We visually inspected the HLA genotype calls by the software, and made corrections as needed. The approved HLA genotype results were exported in Histoimmunogenetics Markup Language (HML) format,^26^ and generated comma separated value (CSV) reports for HLA genotypes, HLA serotypes including Bw4 and Bw6, KIR ligands (C1 and C2) and imputed HLA haplotypes.^27,28^

### Host Sequence Alignment

Low-coverage FASTQ sequences underwent quality control assessment via FastQC v0.11.8 before alt-aware alignment to GRCh38.p12 using BWA-MEM v0.7.17-r1188. Duplicate sequences were marked with MarkDuplicates of the Picard Tools suite v2.21.2. After duplicate marking, base quality score recalibration was performed with Picard Tools’ BaseRecalibrator and high-confidence variant call sets from dbSNP and the 1000 Genomes Project. Quality control metrics, including coverage, were generated with Qualimap BAMQC v2.2.1, Samtools v1.10, and Mosdepth v0.2.9. Finally, quality control reports for each sample were aggregated using MultiQC v1.9. Reproducible code and steps are available at Protocols.io doi: XXX (https://www.protocols.io/private/8CFBD1AD8FE611EA815E0A58A9FEAC2A)

### Variant Calling, Imputation, PCA, Kinship

BAM files were used for an initial calling with bcftools v1.9 mpileup^29^. To account for the low-coverage sequencing we used the GLIMPSE algorithm v1.0 for imputation^3^. Briefly, this algorithm uses a reference set of haplotypes (1000 genomes in our case) to compute genotype likelihoods using a Gibbs sampling procedure. The imputed data were filtered for low imputation scores (INFO>0.8), and were then merged with a reference set that contained samples from: (1) the 1000 genomes project, (2) the HGDP, and (3) the SGDP. While merging these data we set minor allele count thresholds of 5 for our data and 20 for the reference set (e.g., MAC>4 using bcftools), and a stringent call rate threshold (*--geno 0.01* in PLINK2)^30,31^. The resulting VCF was loaded into PLINK2 v2.00a3LM using the following flags: *dosage=DS, --import-dosage-certainty 0.8*. These merged data had 4,111,339 autosomal variants that survived the filters above. PLINK2 was then used for LD pruning (*--indep-pairwise 500 10 0.1*) and PCA (*--maf 0.01 --pca*). We also extracted the kinship matrix of our samples using the King algorithm (*--make-king* in PLINK2)^32^.

## Data Availability

The data referred to in this manuscript is intended for sharing after necessary regulatory approval. At this time, please contact the corresponding authors directly for information on the status of approvals in accordance with the Stanford School of Medicine IRB.

## Acknowledgments

Takeda Pharmaceuticals provided funding support for nucleic acid extraction and library preparation. One Lambda, Inc supplied HLA sequencing reagents and software. The Chan Zuckerberg Initiative and Biohub supported all viral genome sequencing and alignment. This work is supported in part by NIH Common Fund (U240D026629 to EAA and MTW), Arnold O. Beckman independence fellowship (MJS) and American Heart Association (MJS, KS), the National Heart Lung and Blood Institute (K08HL143185 to VNP), The John Taylor Babbitt Foundation (VNP) and Sarnoff Cardiovascular Research Foundation (VNP).Supplemental Material

## Supplemental Material

**Supplemental Figure 1.**
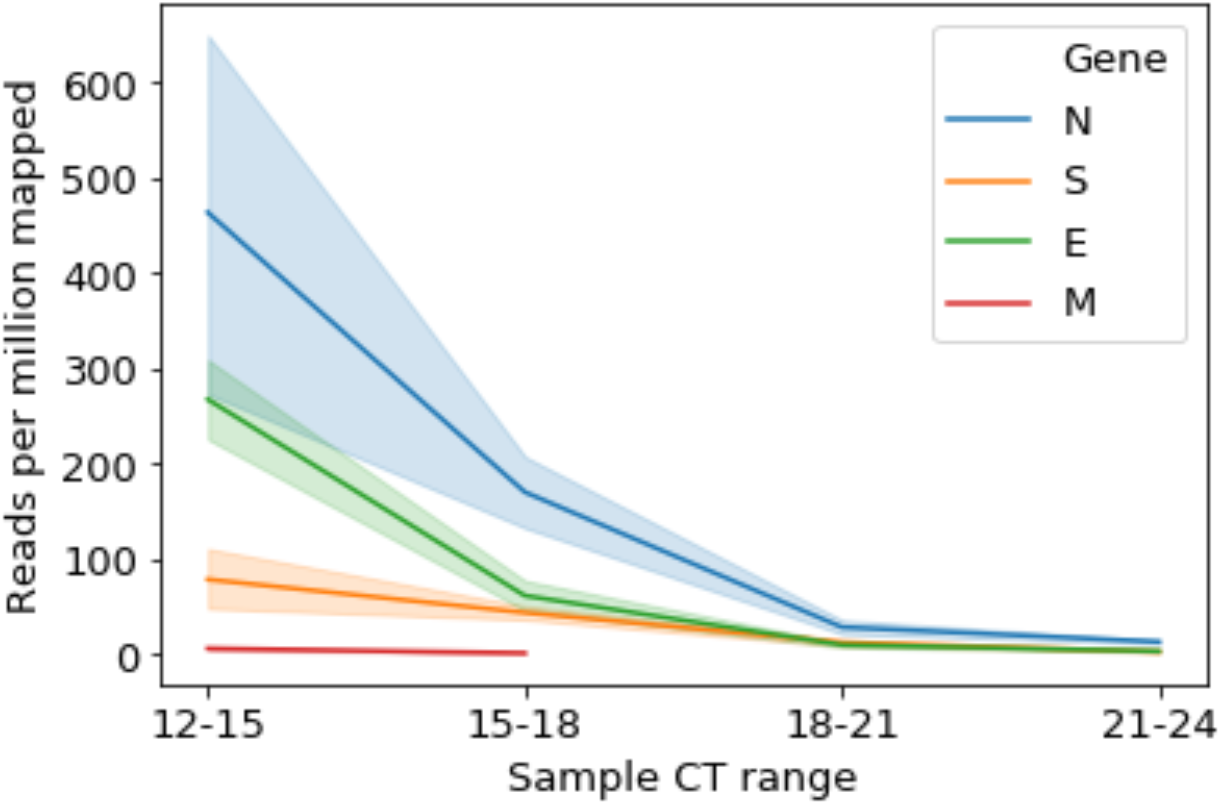
sgRNA expression is correlated with CT range. Patient samples with lower CT values had significantly higher sub-genomic RNA (sgRNA) reads per million mapped (RPM) for genes E (R^2^= 0.23, p=1.4e-17), M (R^2^=0.17, p=1.0e-03), and N (R^2^=0.02, p=5.1e-03). These results show that our method is capable of sequencing SARS-CoV2 viral transcripts as well as genomes. Of note, proteins M and S were observed only in low-CT value samples.

**Supplemental Figure 2.**
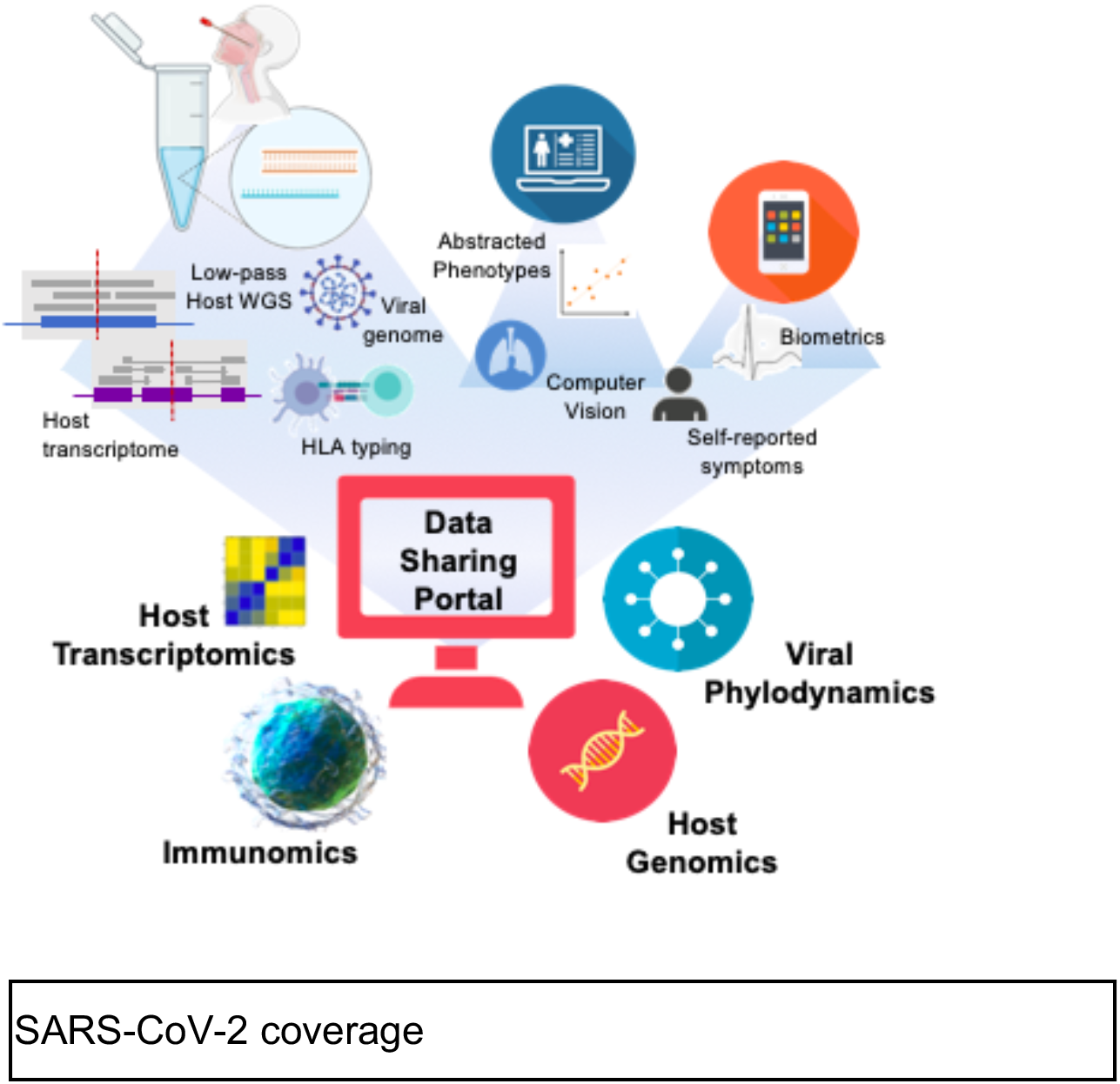
A strong multi-omic foundation for data integration and sharing across global institutions. Using these methods in combination with electronic health record abstraction, and digital medicine, the methods described here builds the foundation for a data repository allowing rapid access to critical data on CoVID19 or any other pandemic via open-source sharing.

**Supplemental Table 1.**
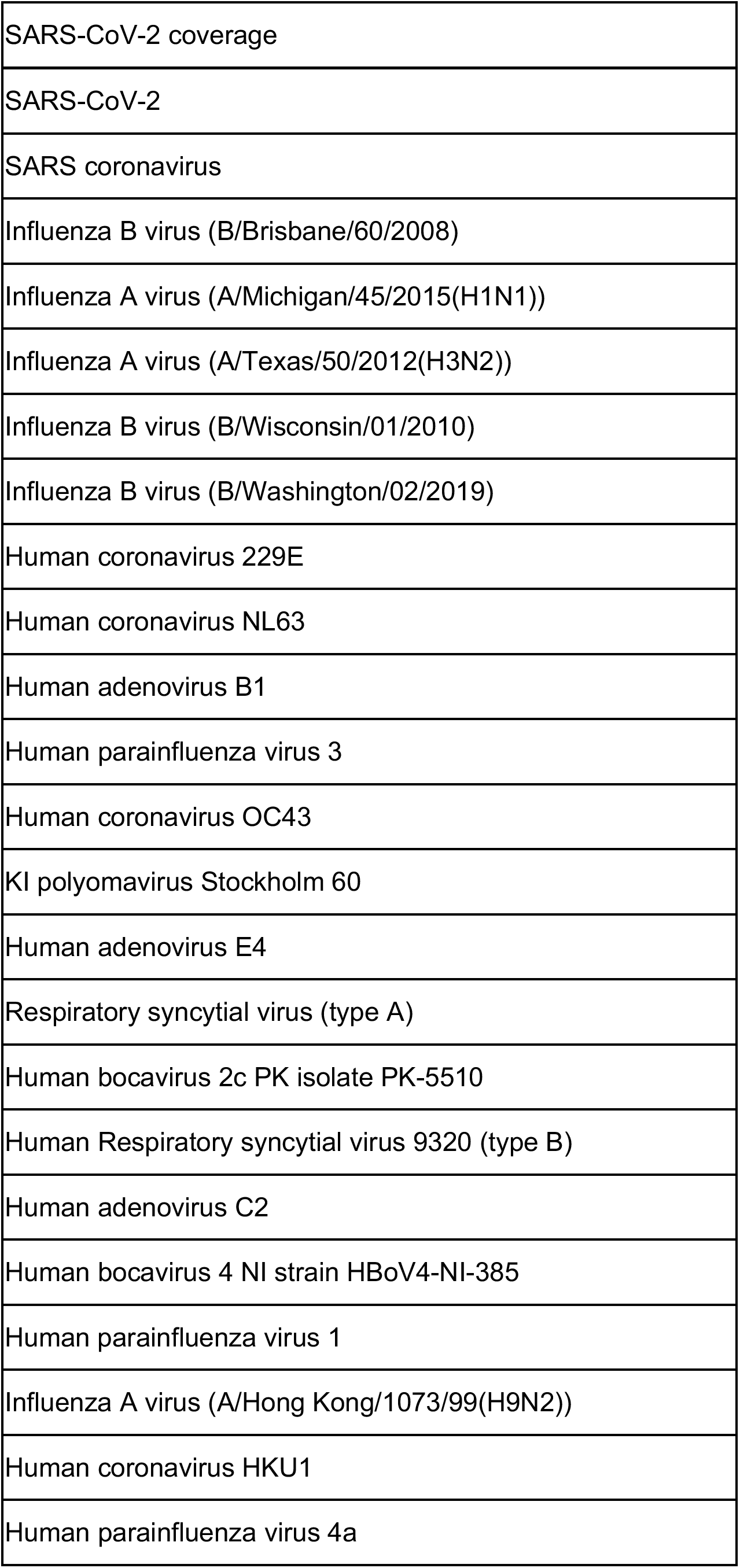
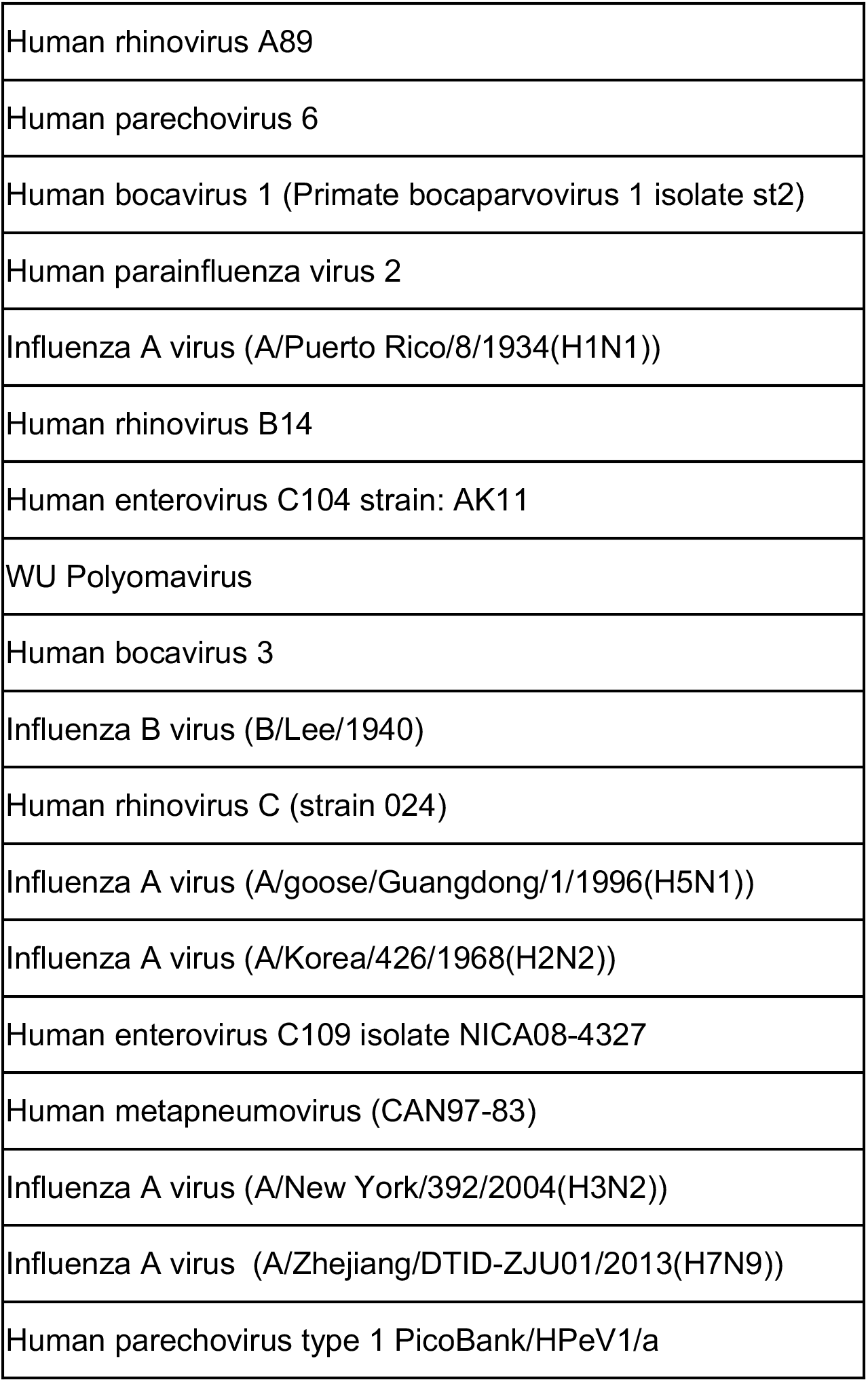
List of viral genomes assessed by alignment using DRAGEN in Illumina BaseSpace.

**Supplementary Table 2.** STAR mapping parameters used for detection of total viral RNA and sgRNA.

| RNA quantification | STAR mapping parameters |
| --- | --- |
| Total viral RNA and sgRNA identification | --outFilterMultimapNmax 20<br>--alignSJoverhangMin 8<br>--outSJfilterOverhangMin 12 12 12 12<br>--outSJfilterCountUniqueMin 1 1 1 1 |
|  | --outSJfilterCountTotalMin 1 1 1 1<br>--outSJfilterDistToOtherSJmin 0 0 0 0<br>--outFilterMismatchNmax 999<br>--outFilterMismatchNoverReadLmax 0.04<br>--scoreGapNoncan -4<br>--scoreGapATAC -4<br>--chimSegmentMin 10<br>--chimOutType WithinBAM SoftClip Junctions<br>--chimScoreJunctionNonGTAG 0<br>--alignSJstitchMismatchNmax -1 -1 -1 -1<br>--alignIntronMin 20<br>--alignIntronMax 1000000<br>--alignMatesGapMax 1000000 |

